# Machine Learning Based Classification of Aggressive and Malignant Renal Tumors from Multimodal Data

**DOI:** 10.1101/2025.02.04.25321687

**Authors:** Mehrnegar Aminy, Tejal Gala, Agnimitra Dasgupta, Steven Y Cen, S J Pawan, Inderbir Gill, Vinay Duddalwar, Assad A Oberai

**Affiliations:** Aerospace and Mechanical Engineering, Viterbi School of Engineering, University of Southern California (USC), Los Angeles, CA, USA; Department of Radiology, Keck School of Medicine of the University of Southern California, Los Angeles, California, USA; Keck School of Medicine, University of Southern California, Los Angeles, CA; Radiomics Lab, Keck School of Medicine of the University of Southern California, Los Angeles, California, USA; USC Institute of Urology and Catherine and. Joseph Aresty Department of Urology Keck School of Medicine, University of Southern California (USC), Los Angeles, CA, USA; Center for Image-Guided Surgery, Focal Therapy and Artificial Intelligence for Prostate Cancer, Keck School of Medicine, University of Southern California, Los Angeles, CA; Alfred E Mann Dept of Biomedical Engineering, USC Viterbi School of Engineering, Los Angeles CA USA 90089; Institute of Urology, University of Southern California, Los Angeles, CA; Dept of Radiology, Los Angeles General Medical Center, Los Angeles, CA

## Abstract

1

**Purpose:** This study aimed to develop and evaluate a machine learning pipeline using multiphase contrast-enhanced CT images and clinical data to classify renal tumors as benign, malignant-indolent, or malignant-aggressive, while assessing the contribution of each data source to the classification.

**Methods:** In this retrospective study, 448 patients (mean age: 60.7±12.6 years, 306 male, 142 female) who underwent nephrectomy and preoperative CECT between June 2008 and July 2018 were included. Tumors were histologically categorized as benign-indolent, malignant-indolent, or malignant-aggressive. Self-supervised feature extraction converted 4-phase CECT images into 512 real-valued features, combined with clinical data and tumor size for classification. Two machine learning classifiers, random forest (RF) and multi-layer perceptron (MLP), were used to predict tumor type. Nested five-fold cross-validation was employed for hyperparameter tuning and model evaluation, and performance was assessed using area under the curve (AUC) analysis.

**Results:** The best-performing models achieved an AUC of 0.90 (95% CI: 0.88–0.93) for classifying indolent versus aggressive tumors and 0.76 (95% CI: 0.71–0.81) for benign versus malignant tumors. Models incorporating tumor size significantly improved classification accuracy. RF classifiers excelled in distinguishing indolent from aggressive tumors, while MLP classifiers performed better for benign versus malignant classification.

**Conclusion:** The machine learning pipeline demonstrated high accuracy in differentiating aggressive from indolent renal tumors, offering valuable prognostic insights for personalized treatment. Tumor size was a critical factor, complementing CECT images and clinical data. These findings highlight the potential of ML techniques in enhancing renal tumor risk stratification.

## 2 Introduction

Kidney cancer is among the top 10 most prevalent cancers in both men and women in the United States and ranks 14th globally. According to the American Cancer Society, approximately 81,610 new cases and 14,390 deaths were projected in the United States in 2024 alone. Prognosis varies widely by stage, with 5-year survival rates of 93% for localized disease, 74% for regional spread, and just 17% for metastatic disease.

Renal tumors present a significant clinical challenge as they are frequently asymptomatic and discovered incidentally during imaging studies(1–4). The increased utilization and availability of cross-sectional imaging, such as computed tomography and magnetic resonance imaging, have led to a stage migration, with renal masses now being identified at smaller sizes and earlier stages before symptoms suggest advanced disease(1, 2, 4, 5). Contrast-enhanced CT (CECT) has become the cornerstone of renal mass evaluation, providing critical information about tumor morphology and enhancement patterns(2, 6). While most incidentally-detected renal masses are simple cysts, benign entities, such as angiomyolipomas and oncocytomas, must be distinguished from malignant renal cell carcinomas (RCCs), which include clear cell, papillary, and chromophobe subtypes(2, 5, 7–10). RCC is the most lethal urologic malignancy, with over 15% of patients presenting with metastatic disease at diagnosis(2, 11, 12). Conventional diagnosis relies on visual interpretation of imaging, which is subject to limitations; accurate qualitative analysis of CECT images is hindered by interobserver variability, intra- and intertumoral heterogeneity, and challenges in comparing contrast differences across imaging phases(2, 8–10).

While tumor size has been shown to correlate with the likelihood of malignancy, size alone, as well as benignity and malignancy categorization, are insufficient to guide modern clinical decision-making(3, 13–16). For example, small renal masses (SRMs), defined as tumors ≤4 cm, now account for up to 66% of new RCC diagnoses, and they are generally categorized into three groups: benign tumors, aggressive cancers, and indolent cancers(4). Although 70–80% of SRMs are malignant, many exhibit indolent behavior over time, as demonstrated in active surveillance series(15, 17). Consequently, not all malignant SRMs require immediate treatment, and indolent malignant tumors can often be managed with active surveillance(4, 15, 18, 19). Management options for SRMs, including surgical resection, thermal ablation, and active surveillance, must be carefully chosen to avoid overtreatment of indolent tumors or undertreatment of aggressive ones(4, 18, 19). Bhindi et al. proposed a framework that stratifies renal masses into histologically indolent and aggressive subgroups based on radiographic size and gender, offering a foundation for individualized treatment decisions(15). However, distinguishing between aggressive and indolent disease remains challenging, even when imaging findings are supplemented with renal mass biopsy data(4, 15, 20).

Improved prognostic tools are needed to enhance risk stratification and provide tailored treatments based on oncologic risk and overall health(4, 15, 21). Advances in machine learning and artificial intelligence offer opportunities to leverage radiographic and clinical data for more precise tumor characterization and behavior prediction(22). While several studies use these tools to classify renal tumors as malignant or benign(23–30), few address their classification as aggressive or indolent(31, 32). In the present study, we fill this gap and quantify the extent to which various data sources, including CECT images, clinical markers, and tumor size contribute to this classification.

In this study, we develop a novel classification pipeline to identify renal tumor aggressiveness using multiphase CECT images and clinical features. Self-supervised feature extraction transforms the CECT images into real-valued features(33), which are then combined with tumor size and clinical data in tabular form for classification using a multi-layer perceptron (MLP) and a random forest (RF) algorithm. Tumors are categorized as benign and indolent, malignant and indolent, or malignant and aggressive. These classifications determine whether a tumor is benign or malignant and, if malignant, whether it exhibits indolent or aggressive behavior.

## 3 Materials and Methods

The overall classification pipeline is presented in Figure 1. The input comprises 4-phase CECT images, clinical features, and tumor size measured using the CECT images. In the first step, images are transformed to real-valued features of dimension 512 using self-supervised feature extraction (Sim-CLR). Once this is accomplished, all input data is in tabular form and can be concatenated and used in a classifier. Two types of classifiers, one based on a multi-layer perceptron (MLP) and the other on a random forest (RF) algorithm are considered. The effect of dimensionality reduction in the input features is also considered. Different permutations of the input data features are used in the MLP and RF classifiers, and each tumor is classified into one of three categories: benign and indolent, malignant and indolent, and malignant and aggressive. Finally, these classifications are combined to determine whether a given tumor is benign or malignant and indolent or aggressive.

**Figure 1.**
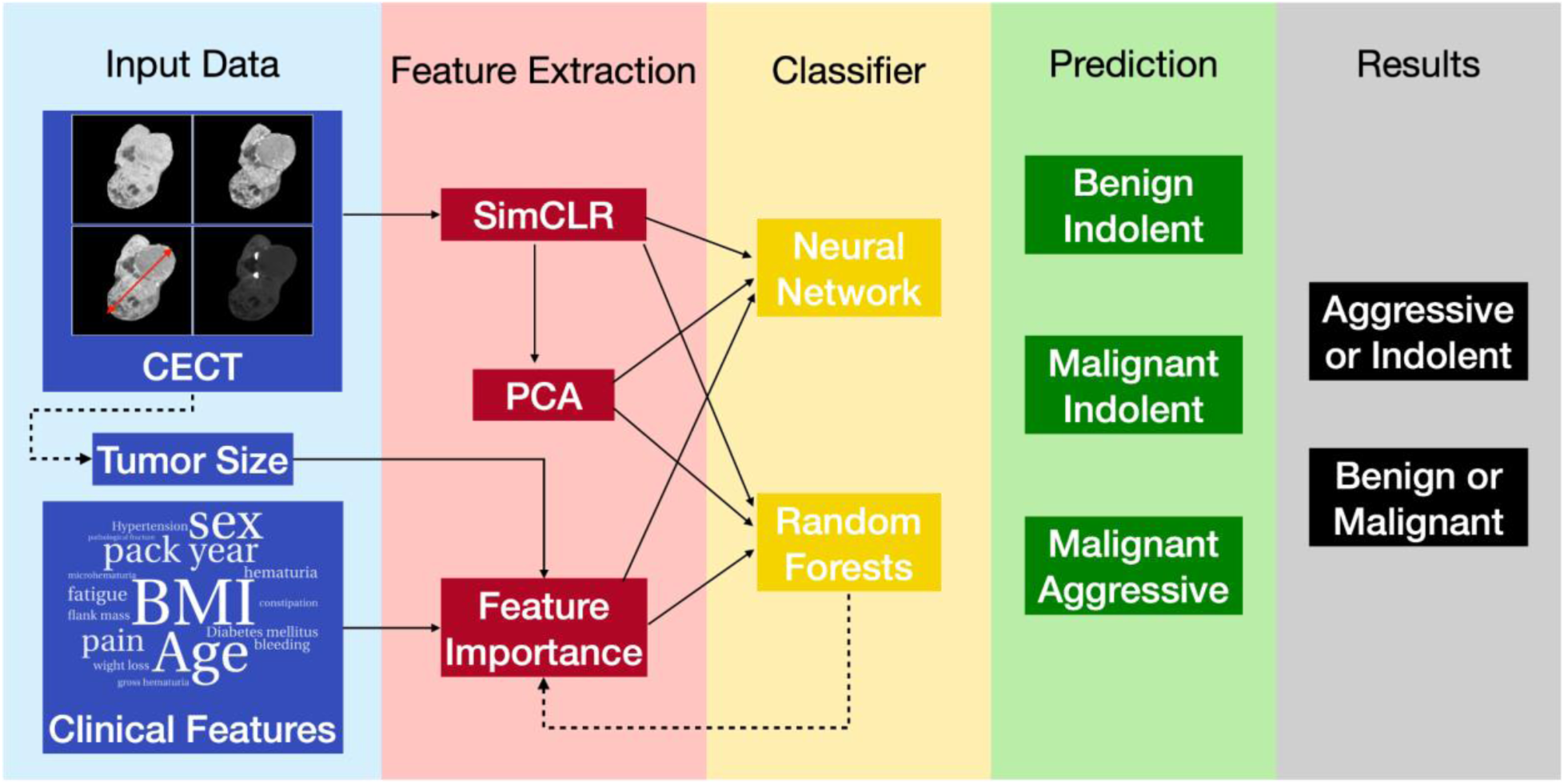
Schematic representation of the pipeline.

### 3.1 Patient Data

This IRB-approved study retrospectively included 448 patients with renal masses at Keck Medical Center of USC who underwent partial or radical nephrectomy and preoperative 4-phase CECT of the abdomen and pelvis between June 2008 and July 2018(Table 1). Post-resection, standard-of-care pathologic evaluation was performed by experienced genitourinary pathologists. The subjects used in this study overlap with those of previous studies(10, 34).

**Table 1.**
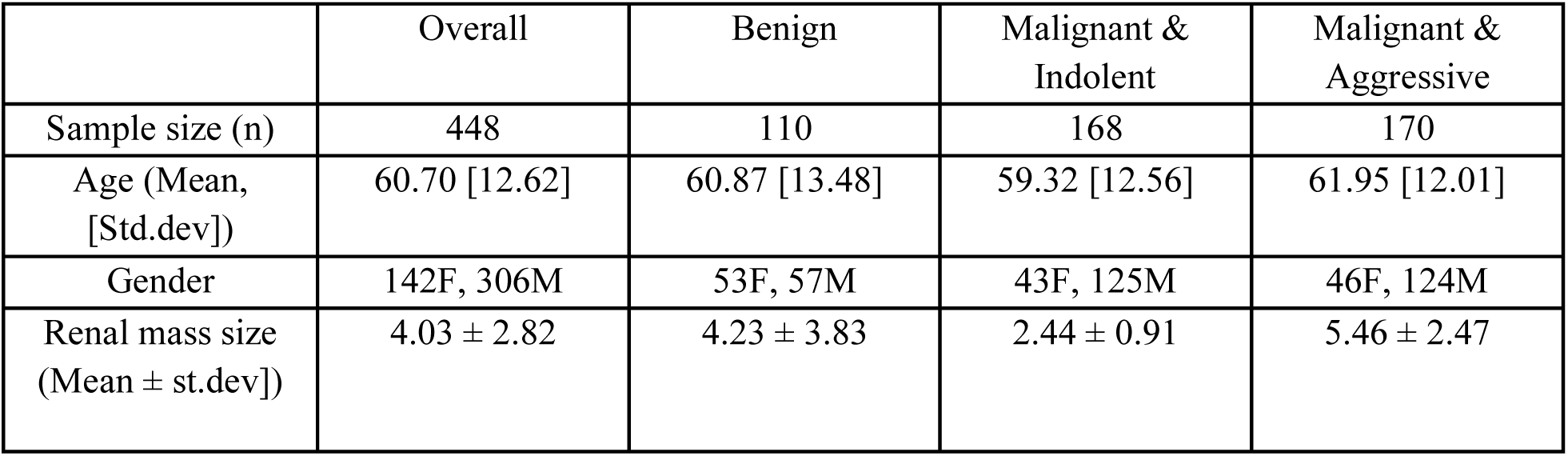
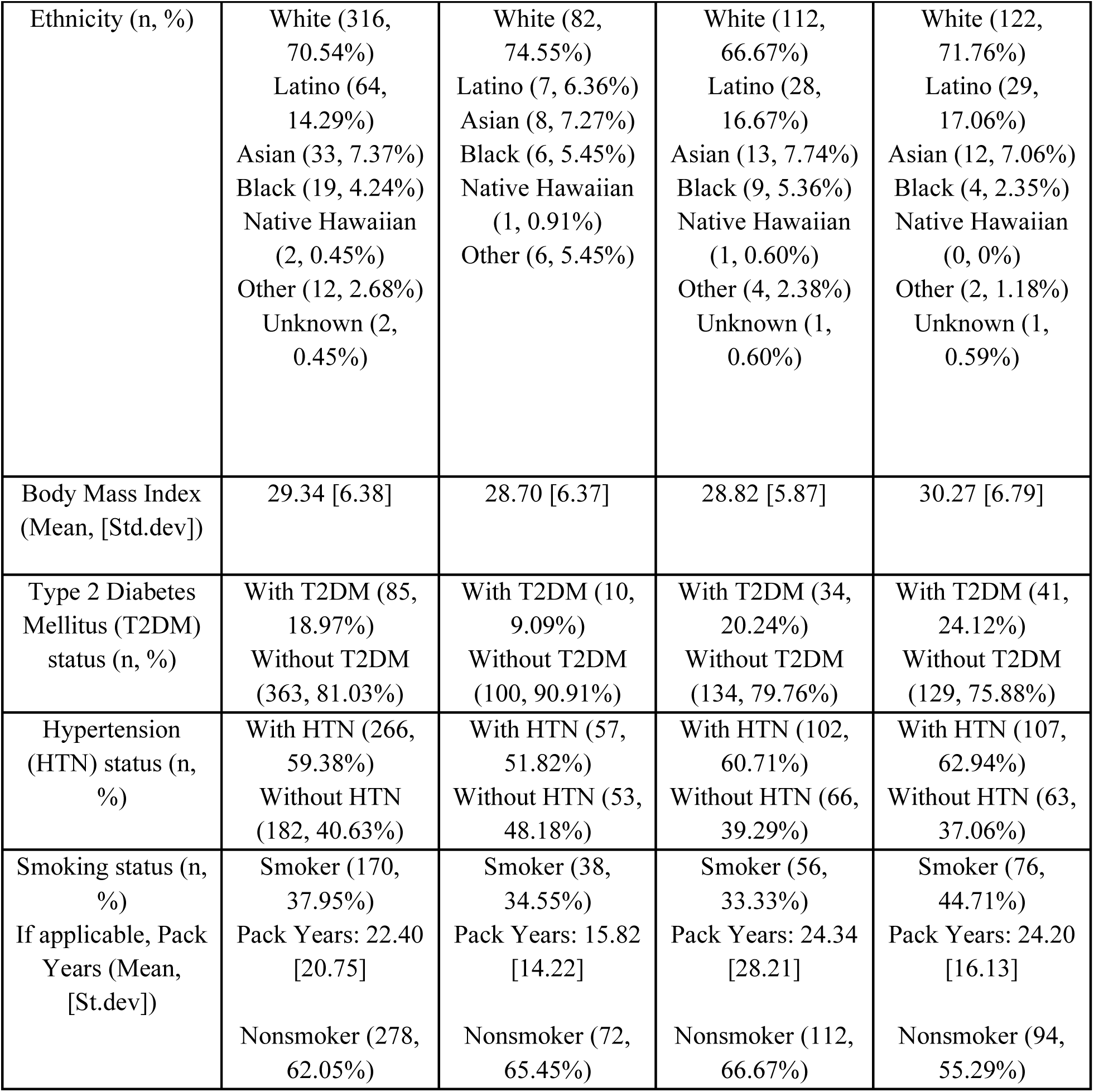
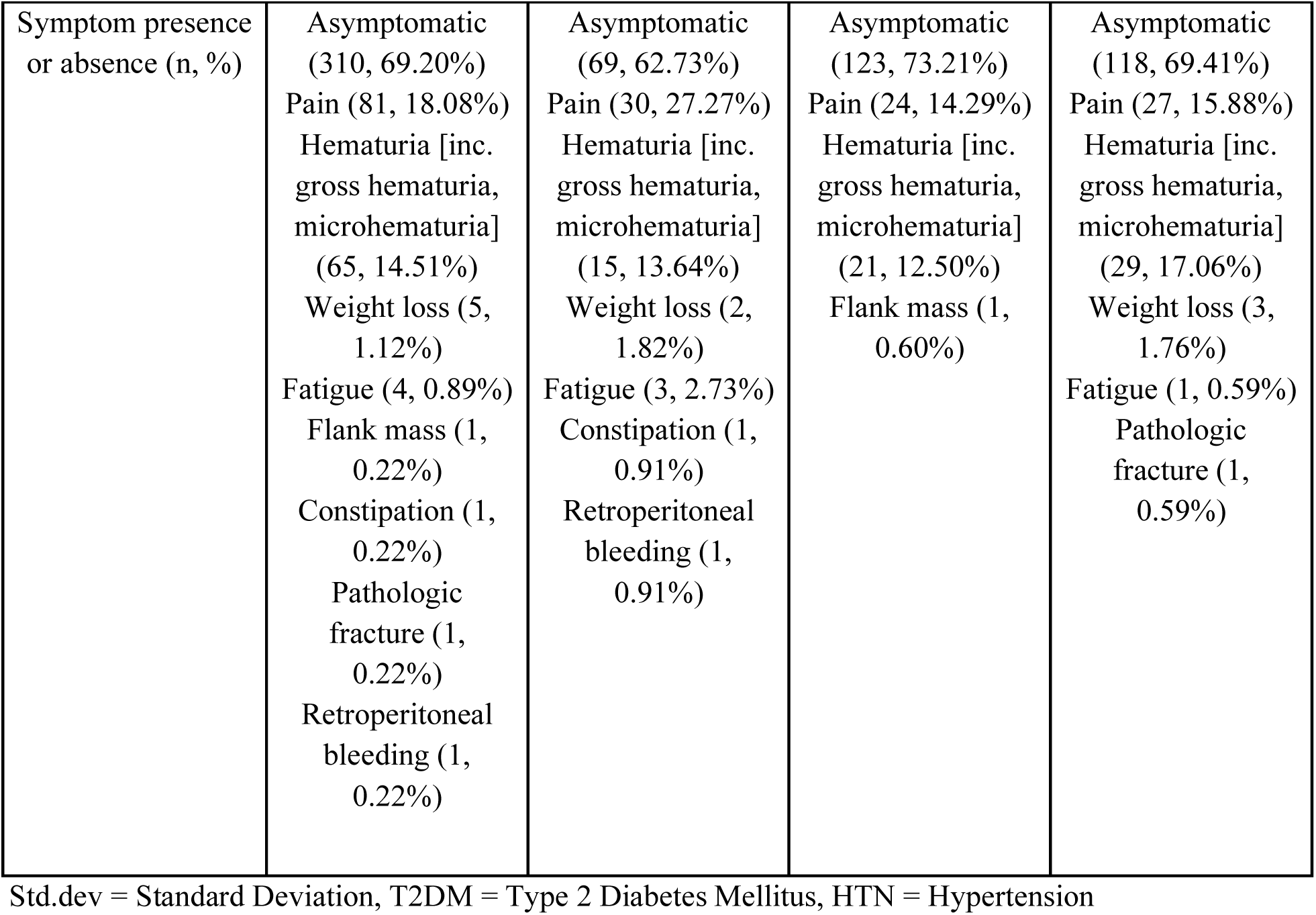
Description of study population.

Image data was deidentified in MATLAB by assigning anonymous study IDs and stored on a secure, password-protected server. Using Synapse 3D (Fujifilm) and blinded to pathologic diagnoses, two senior radiologists-in-training manually segmented renal tumors as three-dimensional regions of interest(34, 35). Images were resampled to a median spacing using nearest-neighbor interpolation, min-max normalized, and center cropped. For each phase, the axial slice passing through the centroid was extracted from the 3D image to create a 2D image with 256x256 pixels. A radiologist with 20 years of abdominal imaging experience verified segmentation accuracy. The three radiologists independently segmented margins for 15 subjects, and reliability was assessed using 2-way mixed intraclass correlation with absolute agreement.

Tumor size was determined as the largest diameter of the tumor across the imaging plane, measured in the nephrographic phase. Clinical features include demographic data, past medical history, and symptomatic data. The demographic data includes body mass index (BMI), age, and sex; past medical history includes the status for diabetes mellitus (DM), hypertension (HTN), and smoking measured in pack-years. Symptomatic data includes gross hematuria, microhematuria, fatigue, weight loss, bleeding, pathologic fracture, constipation, and flank mass. Integer- and real-valued data is normalized to lie between (0, 1), and features like DM and HTN status are represented using binary encoding.

Samples were defined as benign, malignant and indolent, or malignant and aggressive based on histological classification as outlined by Bhindi et al.(15) and shown in Table S1. Benign renal masses include oncocytoma, papillary and metanephric adenomas, and non-epithelioid angiomyolipoma. Malignant indolent tumors encompass low-grade ccRCC and papillary RCC, chromophobe RCC, clear-cell papillary RCC, mucinous tubular and spindle cell RCC, succinyl dehydrogenase-deficient RCC, epithelioid angiomyolipoma, and tubulocystic RCC regardless of grade. Tumors were defined as aggressive if they exhibit sarcomatoid differentiation or coagulative necrosis, except in cases of low-grade (International Society of Urological Pathology grade 1–2) papillary RCC. Malignant aggressive tumors include high-grade ccRCC and papillary RCC, collecting duct RCC, translocation-associated RCC, hereditary leiomyomatosis RCC, unclassified RCC, and other malignant non-RCC tumors.

### 3.2 Feature extraction

Images were mapped into real-valued features which were then treated in the same way as other tabular data. This was accomplished by converting each 256x256x4 CECT image into a vector of dimension 512 using contrastive learning. In this approach, a class of similar images is created by applying transformations like rotation, scaling, and blurring to a given image, and a class of contrastive images is created by using two distinct images from the data set. Thereafter, a classifier is trained to distinguish between images in the two classes, and the last-but-one layer (latent layer) of the classifier is retained as a low-dimensional feature space for the image. Once the classifier is trained, any new image can be passed through this network, and the value of the latent layer is recovered as a feature embedding of that image. This approach is commonly referred to as SimCLR(33), and it has been shown to outperform other techniques for extracting features from images(33). For details, please refer to Appendix A.

### 3.3 Dimension Reduction

The SimCLR features represent a dimension reduction approach that transforms 256x256x4 = 262,144 values for each image to 512 features. Further reduction was achieved by applying principal component analysis (PCA) to the SimCLR features to identify the informative directions in our dataset. It was observed that the first 20 principal components accounted for 98% of the total variance in the data and were therefore used as a low-dimensional representation of each image.

The number of clinical features was reduced from 20 to 9 by applying the permutation feature importance technique within the RF classifier to find the most important clinical features.

### 3.4 Classification

Any tumor can be uniquely categorized into one of three categories: “benign and indolent”, “malignant and indolent” and “malignant and aggressive”. Two types of classifiers wre developed for this three-class classification problem: random forests (RFs) and multi-layer perceptrons (MLPs). The Gini index was used to train the RF network and cross-entropy loss was used for the MLP. The network architecture for all the models is relatively simple and is described in Appendix B. For the MLP models, a learning rate of 3e-4 and L2 regularization factor of 8e-4 were used and the parameters were initialized using the default PyTorch initialization.

Once the category for a tumor was determined, it was determined whether the tumor is malignant or benign and aggressive or indolent as follows. Tumors in the “malignant and indolent” or “malignant and aggressive” categories were classified as malignant, and those in the “benign and indolent” category were classified as benign. Similarly, tumors in the “benign and indolent“ or “malignant and indolent” categories were classified as indolent, while those in the “malignant and aggressive” category were classified as aggressive.

Nested five-fold cross-validation was used for hyperparameter tuning and model evaluation, and the area under the curve (AUC) for the receiver operating characteristic curve was reported for each model (see Appendix C for details).

## 4 Results

In Table 2 and Figure 2, we present results for all models. Models 1-3 use a single source of input data. Models 4 and 5 use clinical features and tumor size. Models 6 to 9 use all sources of input data.

**Figure 2.**
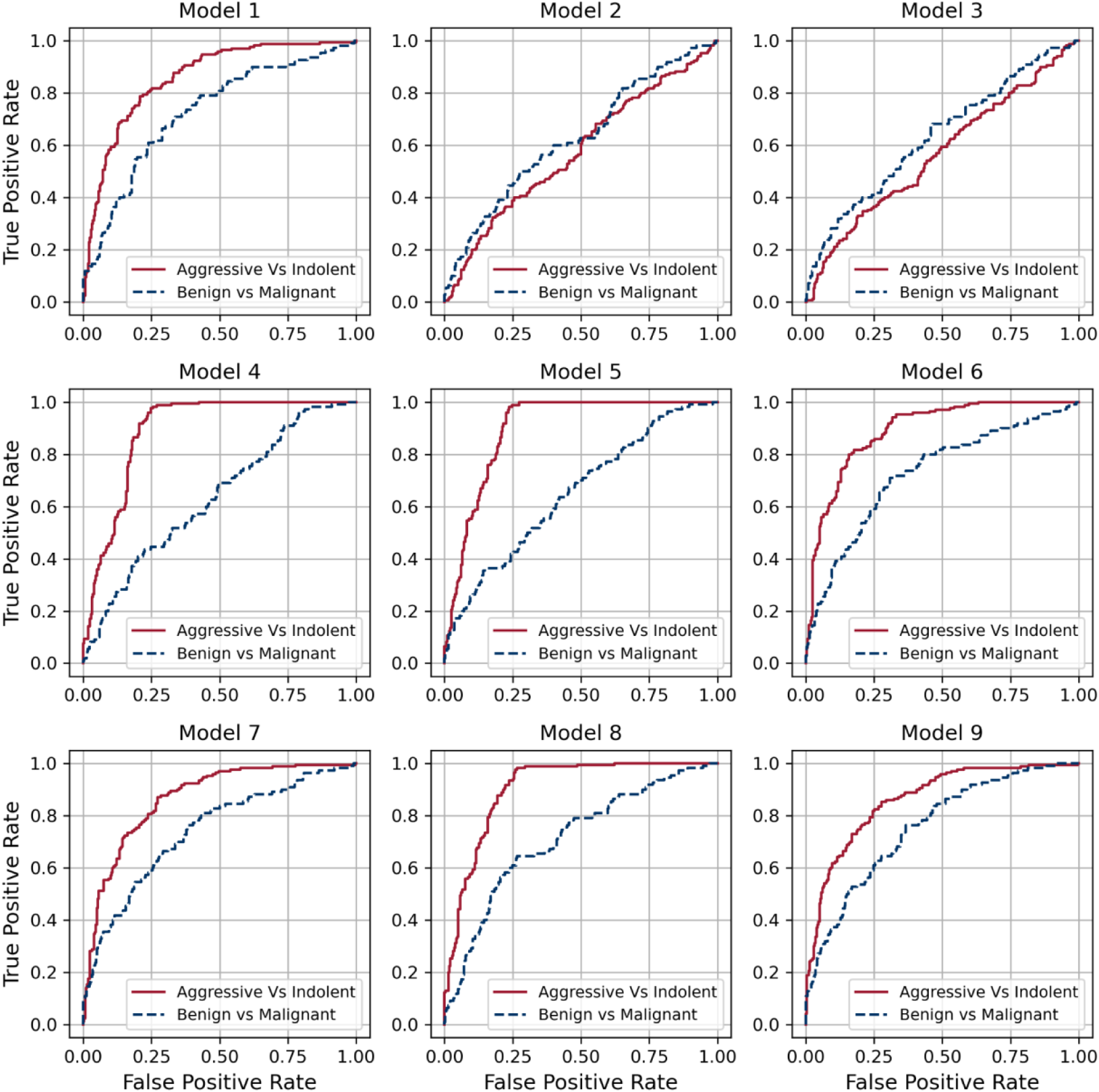
Receiver Operating Characteristic (ROC) curves for Aggressive vs. Indolent and Benign vs. Malignant classifications for all models.

**Table 2.** AUC and 95% confidence interval for different classification models. Models are characterized by their inputs and architecture.

| Model Number | Model Input | Architecture | Aggressive vs Indolent AUC and 95% CI | Malignant vs Benign AUC and 95% CI |
| --- | --- | --- | --- | --- |
| 1 | Images | MLP | 0.86<br>(0.82, 0.89) | 0.73<br>(0.67, 0.78) |
| 2 | Clinical Features (all) | RF | 0.58<br>(0.52, 0.63) | 0.63<br>(0.57, 0.69) |
| 3 | Clinical Features (9) | RF | 0.57<br>(0.51, 0.62) | 0.63<br>(0.57, 0.70) |
| 4 | Clinical Features (all) + Tumor Size (CT) | RF | 0.89<br>(0.86, 0.92) | 0.63<br>(0.57, 0.69) |
| 5 | Clinical Features (9) + Tumor Size (CT) | RF | 0.90<br>(0.87, 0.93) | 0.65<br>(0.59, 0.71) |
| 6 | Images + Clinical Features (9) + Tumor Size (CT) | RF | 0.89<br>(0.86, 0.92) | 0.73<br>(0.67, 0.78) |
| 7 | Images + Clinical Features (9) + Tumor Size (CT) | MLP | 0.87<br>(0.83, 0.90) | 0.74<br>(0.68, 0.79) |
| 8 | Images (PCA) + Clinical Features (9) + Tumor Size (CT) | RF | 0.90<br>(0.88, 0.93) | 0.71<br>(0.66, 0.77) |
| 9 | Images (PCA) + Clinical Features (9) + Tumor Size (CT) | MLP | 0.86<br>(0.83, 0.90) | 0.76<br>(0.71, 0.81) |

CI = Confidence Interval

Model 1 is an MLP that uses image embeddings to perform the classification. It has acceptable performance for both aggressive versus indolent and malignant versus benign classifications with AUCs of 0.86 and 0.73, respectively. Model 2 uses only clinical features as input and underperforms for both types of classifications.

In Fig. 3, using the results of Model 2, we present the relative feature importance values for the clinical features. Thereafter, in Model 3 we use the nine most important clinical features and note that its performance is comparable to that of Model 2.

**Figure 3.**
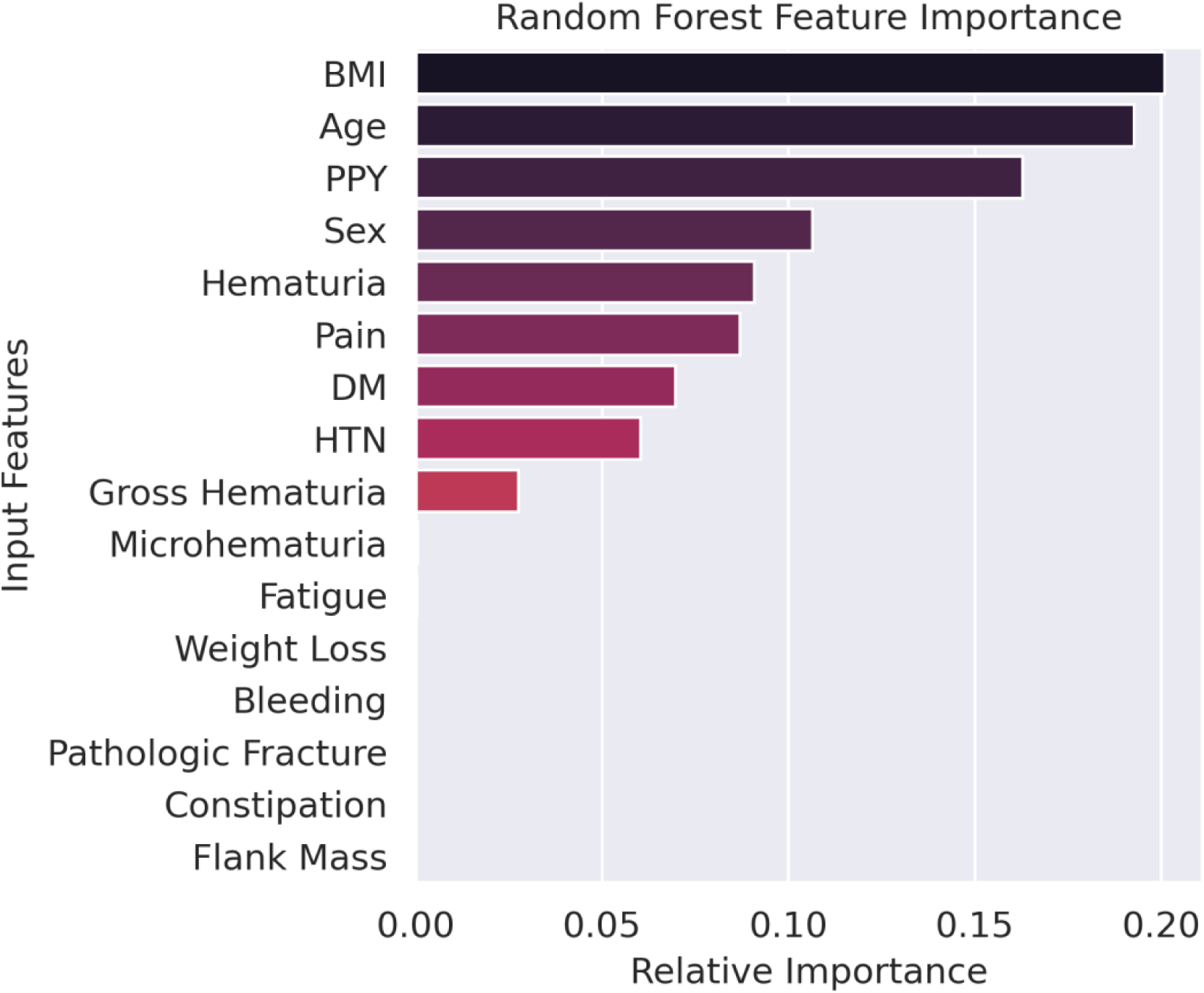
Permutation relative feature importance for clinical data as determined by the random forest model. BMI = Body Mass Index, PPY = Pack Year, DM = Diabetes Mellitus, HTN = Hypertension

In Model 4, we use clinical features, and the tumor size determined from CT images as input and observe that its performance for aggressive vs indolent classification is noticeably better than models that only use clinical features (Models 2 and 3). However, its performance in classifying malignant vs benign lesions is not as good as the model that uses images as input (Model 1). Model 5 is similar to Model 4; however, it uses the reduced set of 9 clinical features. We observe that its performance is like that of Model 4.

Models 6 and 7 use all sources of input data and differ from each other in the architecture of the classifier. Model 6 uses an RF classifier, while Model 7 uses an MLP. We observe that the RF model (Model 6) performs better on the aggressive versus indolent classification task, while the MLP model (Model 7) performs better on the malignant versus benign classification task.

Models 8 and 9 use the first 20 principal components of the image embeddings as input. We note that working with this reduced set does not significantly affect the performance of the models and that the change in AUCs between Models 6 and 8 and Models 7 and 9 is less than or equal to 0.02 points.

### 4.1 Statistical Analysis

In Table 3, we report the difference in the AUC for the aggressive versus indolent classification between pairs of models and the 95% confidence interval (CI) for this difference. The value reported in the (i, j) cell of this matrix is equal to AUC(Model j) - AUC (Model i), and the corresponding 95% CI is derived from Delong’s Z test. Thus, the model considered in column j is superior to the model in row i if this value is positive. Further, this assertion is statistically significant when the 95% CI does not include zero. For example, Model 5 is statistically superior to Model 3 since the difference in the AUC is 0.33, and the 95% CI of this difference does not include zero.

**Table 3.** Difference in the AUC between models for classifying lesions as aggressive or indolent. For column j, and row i, each cell in the table reports the difference between the AUC of Model i and Model j. It also contains (in parenthesis) the 95% confidence interval (CI) of this difference.

| Model Number |  | 1 | 2 | 3 | 4 | 5 | 6 | 7 | 8 | 9 |
| --- | --- | --- | --- | --- | --- | --- | --- | --- | --- | --- |
|  | AUC | 0.86 | 0.58 | 0.57 | 0.89 | 0.90 | 0.89 | 0.87 | 0.90 | 0.86 |
| 1 | 0.86 | - | -0.28<br>(-0.35, -0.22) | -0.29<br>(-0.36, -0.23) | 0.03<br>(-0.01, 0.07) | 0.04<br>(0, 0.08) | 0.03<br>(0.02, 0.05) | 0.01<br>(0, 0.02) | 0.05<br>(0.02, 0.07) | 0<br>(-0.01, 0.02) |
| 2 | 0.58 | 0.28<br>(0.22, 0.35) | - | -0.01<br>(-0.03, 0.01) | 0.31<br>(0.26, 0.37) | 0.32<br>(0.27, 0.37) | 0.31<br>(0.25, 0.38) | 0.29<br>(0.23, 0.35) | 0.33<br>(0.27, 0.39) | 0.29<br>(0.22, 0.35) |
| 3 | 0.57 | 0.29<br>(0.23, 0.36) | 0.01<br>(-0.01, 0.03) | - | 0.32<br>(0.27, 0.38) | 0.33<br>(0.28, 0.39) | 0.32<br>(0.26, 0.39) | 0.30<br>(0.24, 0.36) | 0.34<br>(0.28, 0.4) | 0.30<br>(0.23, 0.36) |
| 4 | 0.89 | -0.03<br>(-0.07, 0.01) | -0.31<br>(-0.37, -0.26) | -0.32<br>(-0.38, -0.27) | - | 0.01<br>(-0.01, 0.02) | 0<br>(-0.03, 0.03) | -0.02<br>(-0.06, 0.01) | 0.01<br>(-0.01, 0.04) | -0.03<br>(-0.07, 0.01) |
| 5 | 0.90 | -0.04<br>(-0.08, 0) | -0.32<br>(-0.37, -0.27) | -0.33<br>(-0.39, -0.28) | -0.01<br>(-0.02, 0.01) | - | -0.01<br>(-0.04, 0.03) | -0.03<br>(-0.07, 0) | 0.01<br>(-0.02, 0.03) | -0.04<br>(-0.07, 0) |
| 6 | 0.89 | -0.03<br>(-0.05, -0.02) | -0.31<br>(-0.38, -0.25) | -0.32<br>(-0.39, -0.26) | 0<br>(-0.03, 0.03) | 0.01<br>(-0.03, 0.04) | - | -0.03<br>(-0.04, -0.01) | 0.01<br>(-0.01, 0.03) | -0.03<br>(-0.05, -0.01) |
| 7 | 0.87 | -0.01<br>(-0.02, 0) | -0.29<br>(-0.35, -0.23) | -0.30<br>(-0.36, -0.24) | 0.02<br>(-0.01, 0.06) | 0.03<br>(0, 0.07) | 0.03<br>(0.01, 0.04) | - | 0.04<br>(0.01, 0.06) | 0<br>(-0.02, 0.01) |
| 8 | 0.90 | -0.05<br>(-0.07, -0.02) | -0.33<br>(-0.39, -0.27) | -0.34<br>(-0.4, -0.28) | -0.01<br>(-0.04, 0.01) | -0.01<br>(-0.03, 0.02) | -0.01<br>(-0.03, 0.01) | -0.04<br>(-0.06, -0.01) | - | -0.04<br>(-0.07, -0.02) |
| 9 | 0.86 | 0<br>(-0.02, 0.01) | -0.29<br>(-0.35, -0.22) | -0.30<br>(-0.36, -0.23) | 0.03<br>(-0.01, 0.07) | 0.03<br>(0.01, 0.05) | 0.03<br>(0.01, 0.05) | 0<br>(-0.01, 0.02) | 0.04<br>(0.02, 0.07) | - |

AUC = Area Under the Curve

From Table 3 we observe that Models 5, 6, and 8 are statistically superior to all other models for the aggressive versus indolent classification. We also observe that the difference among these three models is very small, and we may claim that their performance models is equivalent.

In Table 4, we report the corresponding values for the difference in the AUC for the malignant versus benign classification. We conclude that Model 9 is statistically better than all models except Models 6 and 7. The 95% CI of the difference in the AUC between Model 9 and Model 6 and Model 9 and Model 7 includes zero; therefore, the performance of these models may be considered equivalent. However, when we enumerate the number of models that Models 6, 7, and 9, are superior to, we conclude that this number is 4 for Models 6 and 7, and 6 for Model 9. Based on this comparison, we conclude that Model 9 is preferable to Models 6 and 7.

**Table 4.** Difference in the AUC between models for classifying lesions as malignant or benign. For column j, and row i, each cell in the table reports the difference between the AUC of Model i and Model j. It also contains (in parenthesis) the 95% confidence interval (CI) of this difference.

| Model Number |  | 1 | 2 | 3 | 4 | 5 | 6 | 7 | 8 | 9 |
| --- | --- | --- | --- | --- | --- | --- | --- | --- | --- | --- |
|  | AUC | 0.73 | 0.63 | 0.63 | 0.63 | 0.65 | 0.73 | 0.74 | 0.71 | 0.76 |
| 1 | 0.73 | - | -0.10<br>(-0.18, -0.02) | -0.09<br>(-0.17, -0.01) | -0.09<br>(-0.17, -0.02) | -0.08<br>(-0.15, 0) | 0<br>(-0.04, 0.04) | 0.01<br>(-0.01, 0.03) | -0.01<br>(-0.05, 0.03) | 0.03<br>(0, 0.06) |
| 2 | 0.63 | 0.10<br>(0.02, 0.18) | - | 0.01<br>(-0.02, 0.03) | 0.01<br>(-0.03, 0.04) | 0.02<br>(-0.01, 0.05) | 0.1<br>(0.02, 0.18) | 0.11<br>(0.04, 0.18) | 0.09<br>(0.01, 0.16) | 0.13<br>(0.06, 0.19) |
| 3 | 0.63 | 0.09<br>(0.01, 0.17) | -0.01<br>(-0.03, 0.02) | - | 0<br>(-0.04, 0.04) | 0.02<br>(-0.01, 0.04) | 0.09<br>(0.01, 0.17) | 0.11<br>(0.04, 0.17) | 0.08<br>(0.01, 0.15) | 0.12<br>(0.06, 0.19) |
| 4 | 0.63 | 0.09<br>(0.02, 0.17) | -0.01<br>(-0.04, 0.03) | 0<br>(-0.04, 0.04) | - | 0.02<br>(-0.01, 0.04) | 0.09<br>(0.02, 0.17) | 0.11<br>(0.04, 0.17) | 0.08<br>(0.01, 0.15) | 0.12<br>(0.06, 0.18) |
| 5 | 0.65 | 0.08<br>(0, 0.15) | -0.02<br>(-0.05, 0.01) | -0.02<br>(-0.04, 0.01) | -0.02<br>(-0.04, 0.01) | - | 0.08<br>(0, 0.15) | 0.09<br>(0.02, 0.15) | 0.06<br>(0, 0.13) | 0.11<br>(0.05, 0.16) |
| 6 | 0.73 | 0<br>(-0.04, 0.04) | -0.1<br>(-0.18, -0.02) | -0.09<br>(-0.17, -0.01) | -0.09<br>(-0.17, -0.02) | -0.08<br>(-0.15, 0) | - | 0.01<br>(-0.03, 0.05) | -0.01<br>(-0.05, 0.03) | 0.03<br>(-0.01, 0.07) |
| 7 | 0.74 | -0.01<br>(-0.03, 0.01) | -0.11<br>(-0.18, -0.04) | -0.11<br>(-0.17, -0.04) | -0.11<br>(-0.17, -0.04) | -0.09<br>(-0.15, -0.02) | -0.01<br>(-0.05, 0.03) | - | -0.03<br>(-0.06, 0.01) | 0.02<br>(-0.01, 0.04) |
| 8 | 0.71 | 0.01<br>(-0.03, 0.05) | -0.09<br>(-0.16, -0.01) | -0.08<br>(-0.15, -0.01) | -0.08<br>(-0.15, -0.01) | -0.06<br>(-0.13, 0) | 0.01<br>(-0.03, 0.05) | 0.03<br>(-0.01, 0.06) | - | 0.04<br>(0.01, 0.08) |
| 9 | 0.76 | -0.03<br>(-0.06, 0) | -0.13<br>(-0.19, -0.06) | -0.12<br>(-0.19, -0.06) | -0.12<br>(-0.18, -0.06) | -0.11<br>(-0.16, -0.05) | -0.03<br>(-0.07, 0.01) | -0.02<br>(-0.04, 0.01) | -0.04<br>(-0.08, -0.01) | - |

## 5 Discussion

When considering models that utilize all input sources, we observe that the models can classify aggressive versus indolent lesions more accurately than benign versus malignant lesions. The best-performing models for the former (Models 5 and 8) have an AUC of 0.90, whereas the best-performing model (Model 9) for the latter has an AUC of 0.76. The high accuracy in classifying aggressive versus indolent lesions implies that we can provide the physician with a sufficiently accurate assessment so that they may recommend different treatments for patients in these two groups.

When differentiating aggressive lesions from indolent lesions, the knowledge of the size of the lesion plays an important role. This is concluded by comparing the AUC for Models 2 and 3, whose input does not include tumor size information, with all other models, whose input includes tumor size information. In each case, the AUC for Model 2 or 3 is smaller, and this difference is statistically significant.

The judicious reduction in the data dimensions does not influence the performance of the model. The dimension of clinical data is reduced from 20 to 9, and that of image features is reduced from 512 to 20. To assess the effect of the former, we compute the difference between the AUCs of Models 2 and 3 and observe that the 95% CI interval of this difference includes zero, thereby indicating that the difference between these models is not significant. For the latter, we compute the difference in the AUCs between Models 6 and 8, and Models 7 and 9 and once again conclude that the performance of the models with the full and reduced set of features is comparable. The advantage of working with a smaller set of features is two-fold. First, for clinical features, it means that the user collects and curates a smaller dataset. Second, for both clinical and image features, it means that the resulting model has fewer trainable parameters and therefore requires less labeled data to train.

When considering all sources of data and a reduced set of features (Models 8 and 9), the RF-based classifier (Model 8) performs better when classifying aggressive versus indolent lesions, while the MLP-based classifier performs better when classifying malignant versus benign lesions. The difference in the AUCs for these models is significant. While it is difficult to tease out why this is the case, this observation implies that a simple partition of the data space into hyper-rectangles accomplished by the RF algorithm is optimal in distinguishing aggressive and indolent tumors. On the other hand, distinguishing malignant and benign tumors requires a more complex partition which is achieved by an MLP.

Models 5, 6, and 8 performed the best when classifying aggressive versus indolent lesions. Among these, Model 5 is particularly attractive because of its simplicity. Its input comprises only the nine important features, and the tumor size is estimated using a single slice of the CT image. The small number of input parameters also implies that the model is easy to train and does not require much data for robust performance.

For classifying malignant versus benign lesions, Model 9 performed the best. This input to this model is also low-dimensional as it includes 9 clinical features, 20 features derived from CECT images, and a single feature that represents the tumor size. However, generating this input requires a significant amount of data collection and processing. It requires all four phases of the CECT images, computing the SimCLR features for these images, dimension reduction via PCA, and explicit calculation of the tumor size. This underlines the fact that distinguishing malignant and benign lesions is a hard problem that requires data from multiple sources.

This study has several limitations. First, even though the data was collected using multiple scanners at three different hospitals, it belongs to a single center, and this may limit the generalizability of the trained model to a broader population. Second, in the current study, the benign class is the minority class, and this may introduce some bias. Given these factors, we propose that future studies should use larger, multi-institutional, well-balanced datasets to build on our results.

In the long term, the models developed in this study can be used as an assistive tool in clinical practice to diagnose renal masses as aggressive and/or malignant. The ability to distinguish aggressive tumors from indolent tumors can be used as an improved prognostic tool to enhance risk stratification and ensure that each patient receives tailored treatment based on their oncologic risk and overall health.

## Data Availability

All data produced in the present study are available upon reasonable request to the authors

## 6 Acknowledgments

The authors acknowledge the Center for Advanced Research Computing (CARC, carc.usc.edu) at the University of Southern California for providing computing resources that have contributed to the research results reported within this publication.

## Abbreviations

CECT: Contrast-Enhanced Computed Tomography
RCC: Renal Cell Carcinoma
SRM: Small Renal Mass
MLP: Multilayer Perceptron
RF: Random Forest
PCA: Principal Component Analysis
DM: Diabetes Mellitus
HTN: Hypertension
ccRCC: Clear Cell Renal Cell Carcinoma
AUC: Area Under the Curve

## 8 Appendix A

In our application of SimCLR, we use the following augmentations using torchvison.transforms: vertical flip, uniformly sampled rotation from -5 to 5 degrees, scaling with a scaling factor uniformly sampled from an interval of 0.9 to 1.1, Gaussian blur with a kernel size of 25, and additive Gaussian noise (zero mean and standard deviation of 0.1) multiplied with a Bernoulli RV with p = 0.5. For the network, we use ResNet-18 with four input channels for the encoder and a simple MLP with 128 neurons followed by the ReLU activation function for the projection head.

## 9 Appendix B

In this section, we describe the architecture of the models employed in this study. We used Python, scikit-learn, and PyTorch to train and evaluate the models. In the notation used below “+” represents the concatenation of vectors, “-” represents the subsequent layer of a network, RF refers to the random forest algorithm, FC layer (I, J) is a fully connected layer with weights and biases and with an input vector of I components and an output vector of J components, ReLU refers to component-wise ReLU activation, and Softmax refers to the softmax activation.

- Model 1: Image Features (512) - FC Layer (512, 3) - Softmax - Class Probabilities (3).
- Model 2: Clinical Features (16) - RF - Class Probabilities (3).
- Model 3: Clinical Features (9) - RF - Class Probabilities (3).
- Model 4: Clinical Features (16) + Tumor Size (1) - RF - Class Probabilities (3).
- Model 5: Clinical Features (9) + Tumor Size (1) - RF - Class Probabilities (3).
- Model 6: Image Features (512) + Clinical Features (9) + Tumor Size (1) - RF - Class Probabilities (3).
- Model 7: Image Features (512) - FC Layer (512, 10) - ReLU + Clinical Features (9) + Image size (1) - FC Layer (20, 3) - Softmax - Class Probabilities (3).
- Model 8: Image Features (20) + Clinical Features (9) + Image Size (1) - RF - Class Probabilities (3).
- Model 9: Image Features (20) + Clinical Features (9) + Tumor Size (1) - FC Layer (20, 3) - Softmax - Class Probabilities (3).

## 10 Appendix C

We use nested five-fold cross-validation for hyperparameter tuning and model evaluation. This process begins by splitting data into two sets: a training and validation set (80%) and a test set (20%). Next, we perform five-fold cross-validation on the training and validation set to identify the optimal hyperparameters. This involves splitting this set into 5 folds, using each fold for validation while training on the remaining folds. The optimal hyperparameters identified are retained, and the model is trained using the entire training and validation set. Thereafter, we evaluate the trained model on the test set, which remains unseen to the network during the hyperparameter tuning process. We repeat this procedure for five distinct non-overlapping test sets to find probability predictions for each subject. Finally, we use these probabilities to construct a receiver operating characteristic curve and compute the area under curve (AUC) for each model.

**Table S1.**
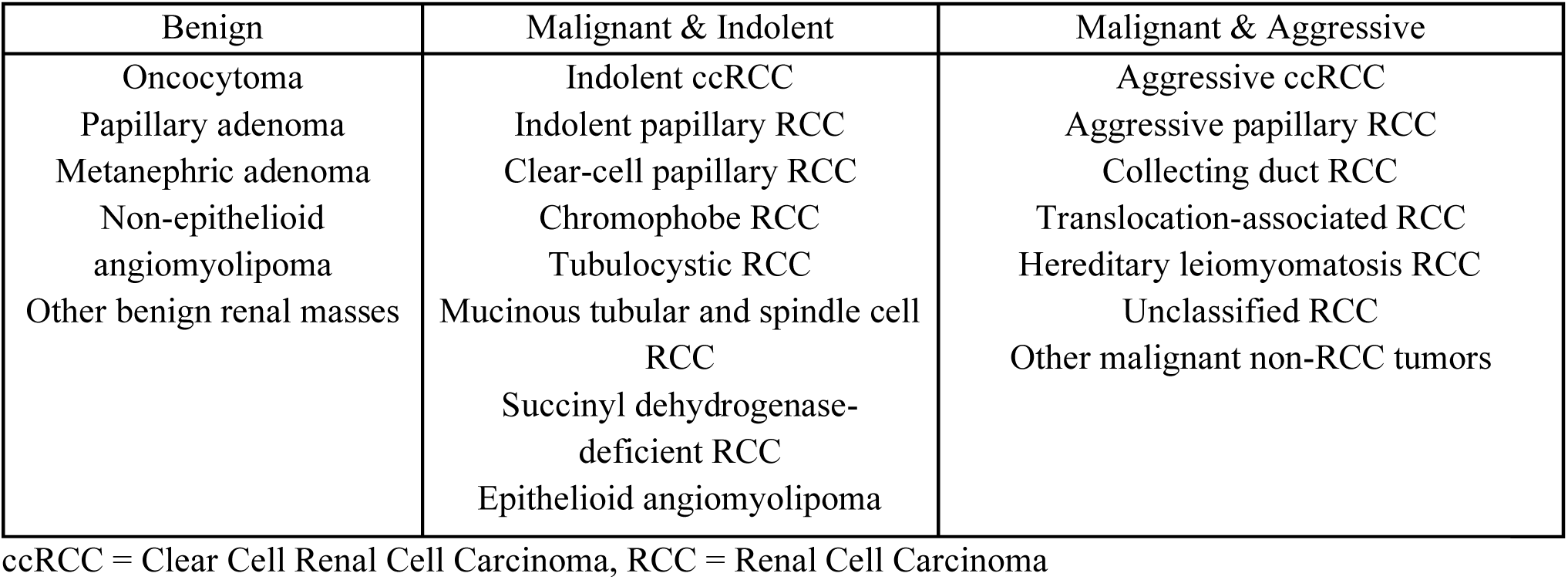
Histologic classification of renal masses.

## Notes

### Competing Interest Statement

The authors have declared no competing interest.

### Funding Statement

This study did not receive any funding.

### Author Declarations

This study was approved by the IRB at the University of Southern California.

